# A fast and sensitive absolute quantification assay for the detection of SARS-CoV□2 peptides using Parallel Reaction Monitoring Mass Spectrometry

**DOI:** 10.1101/2022.03.18.22272462

**Authors:** Akshada Gajbhiye, Atakan Nalbant, Tiaan Heunis, Frances Sidgwick, Andrew Porter, Yusri Taha, Matthias Trost

## Abstract

The on-going SARS-CoV-2 (COVID-19) pandemic has called for an urgent need for rapid and high-throughput methods for mass testing for early detection, prevention and surveillance of the disease. Here, we tested if targeted parallel reaction monitoring (PRM) quantification using high resolution Orbitrap instruments can provide the sensitivity and speed required for a high-throughput method that could be used for clinical diagnosis. Here we report a high-throughput and sensitive PRM-MS assay that enables absolute quantification of SARS-CoV-2 nucleocapsid peptides with short turn-around times. Concatenated peptides (QconCAT) synthesized using isotopically labelled SARS-CoV-2 were used for absolute quantification. We developed a fast and high-throughput S-trap-based sample preparation method, which was then successfully utilized for testing 25 positive and 25 negative heat-inactivated nasopharyngeal swab samples for SARS-CoV-2 detection. The method was able to differentiate between negative and positive patients accurately within its limits of detection. Moreover, extrapolating from the QconCAT absolute quantification, our data show that patients with Ct values as low as 17.5 have NCAP protein amounts of around 7.5 pmol in swab samples. The present high-throughput method could potentially be utilized in specialized clinics as an alternative tool for detection of SARS-CoV-2.

## INTRODUCTION

The on-going human Coronavirus Disease 2019 (COVID-19) pandemic caused and still causes severe global health and economic problems in almost every country in the world. Since the beginning of the pandemic till March 2022, Covid-19 has affected more than 450M people and caused 6 M deaths worldwide [1]. The disease is caused by severe acute respiratory syndrome coronavirus 2 (SARS-CoV-2), a novel and more virulent strain of coronaviruses (CoVs) [2]. This strain of coronavirus is more virulent but similar to two other strains of betacoronaviruses SARS-CoV and Middle East respiratory syndrome (MERS-CoV) [3]. Like other CoVs, the SARS-CoV-2 genome comprises of linear, single-stranded positive-sense RNA which encodes 10 genes, responsible for production of total 26 proteins. Out of the 26 proteins, four structural proteins contribute to ∼80% of the genome. These four structural proteins comprise of a spike glycoprotein (S), which enables viral entry into the mammalian cell by binding to the angiotensin-converting enzyme 2 (ACE2) receptor, a nucleoprotein (N) that provides stability to the viral genome by directly binding to the RNA, an envelope protein (E) and membrane protein (M) that forms the outer layer of the virus [4]. The N and S protein copies per virion are estimated to be approximately 1000 and 300, respectively. The remaining proteins of the genome are required for functions such as proofreading, RNA polymerase, proteases and other supporting proteins for replication of the genome.

The predominant method of testing for individuals infected with SARS-CoV-2 includes real-time quantitative polymerase chain reaction (RT-qPCR), which is usually done on nasopharyngeal (NP) swabs. The RNA is extracted from these swabs and amplified using specific primers, which makes it more specific, sensitive and relatively rapid, deeming it as the gold standard assay for SARS-CoV-2 detection by World Health Organization (WHO). Other rapid tests including Simple Amplification Based Assay (SAMBA) [5], which uses nucleic acid for detection, serological assays such as Lateral Flow Immunoassays (LFA) [6] and enzyme-linked immunosorbent assays (ELISA) have also been developed. These have been implemented owing to the global pressure for identifying the infected individuals with quick turnaround rates. The high demand for RT-qPCR testing has in recent times caused a global shortage of reagents as well as other rapid tests are prone to false-positive and false-negative reporting which might be caused due to inhibition of substances in clinical samples [7]. Thus, at times during the pandemic, complementary alternative assays for detection of SARS-CoV-2 became increasingly urgent to share the burden of immediate mass testing required for management of the pandemic.

In recent years mass spectrometry (MS)-based targeted proteomic approaches have increasingly been implemented in clinical labs due to advancements in the sensitivity and accuracy of instrumentation. An excellent example of this is successful application of Matrix-Assisted Laser Desorption/Ionization Time-Of-Flight mass spectrometry (MALDI-TOF-MS) for characterization of Nucleoprotein (NCAP) and Spike Glycoprotein (SPIKE) from the SARS virus, which caused an outbreak in 2003[8]. Building on this, many MS-based methods have emerged for detection of the SARS-CoV-2 NCAP, and SPIKE proteins based on their tryptic peptides. Most of these methods either have been built directly from specimens infected with high viral loads of SARS-CoV-2; for instance, a study by Gouveia *et al* selected 14 peptides from proteomics of SARS-CoV-2 infected Vero cells [9], or from clinical specimens where enrichment was required for detection of these peptides. Ihling *et al* used gargle solution for detection of SARS-CoV-2 peptides from three Covid-19 positive individuals [10]. In another study by Nikolaev *et al*, peptides from NCAP protein were detected by tandem mass spectrometry using nasopharyngeal epithelial scrapings [11]. Similarly, Singh *et al* [12], Gouveia *et al* [13] and Saadi *et al* [14] utilized nasopharyngeal swabs to establish proof-of-principle studies for detection of SARS-CoV-2 peptides. More recently Renuse *et al* [15] present a LC-MS based method where peptides for NCAP and SPIKE were enriched using immunoaffinity beads. Puyvelde *et al* [16] have attempted to use a QconCAT Stable Isotopic Labeled (SIL) internal standard for improving the efficiency of their MRM-MS assay. Importantly, most of these methods include acetone precipitation-based methods for sample preparation and are either low-throughput or time consuming, thereby lacking the short turn-around time required for a clinical high-throughput assay.

In this study, we report development of a high-throughput liquid chromatography mass spectrometry based parallel reaction monitoring (PRM) assay for detection and absolute quantification of SARS-CoV-2 peptides, which allows viral detection directly from patient samples. We could identify and report patients with high viral load and Ct values up to 22. This method allows detection of SARS-CoV-2 peptides with a turnaround time of ∼2 hrs for each patient starting from sample preparation to reporting the results and up to 200 samples per day.

## MATERIALS AND METHODS

### Chemicals and reagents

Sodium dodecyl sulfate (SDS), >98% pure was purchased from VWR International Limited. Triethylammonium bicarbonate (TEAB) 1 M, and ortho-phosphoric acid 85%, Tris(2-carboxyethyl) phosphine (TCEP) and Iodoacetamide ≥ 98% were purchased from Sigma. Proteomics Grade Trypsin was purchased from Pierce Thermo Fisher Scientific. Acetonitrile, Methanol (LC-MS grade) and Formic Acid (>99.0%, LC-MS, UHPLC-MS) were purchased from Fisher Scientific. Formic acid was obtained from Merck. S-traps were purchased from Protifi. SARS-CoV-2 Spike Glycoprotein (Full-Length) expressed in CHO cells from Native Antigen, Synthetic Sputum from LGC and PolyQuant Cov-MS from PolyQuant were received as part of the Covid Moonshot Consortium. SARS-CoV-2 Nucleoprotein was received from University of Sheffield.

### Patient samples for method validation

Fifty nasopharyngeal swab samples including both positive and negative for Covid-19, were received from the Newcastle Upon Tyne Hospitals NHS Foundation Trust, in 2.5 mL VTM. The samples were stored at -20°C until further processing.

### Sample pre-processing and preparation for MS

The SARS-CoV-2 recombinant protein standards were processed for peptide identification, targeted MS-based proteomic method development, and calibration curve establishment for sensitivity assessment. Each protein standard was processed separately by suspension trapping (S-Trap), with minor modifications as previously described [17,18], for method development and peptide identification. Modifications included a single step reduction and alkylation with 10 mM TCEP and IAA, respectively, and digestion at 47 °C for 1 hour using a 1:10 trypsin: protein ratio. The method was further optimized to exclude the reduction and alkylation step, as well as sample elution, to achieve a faster and high-throughput method discussed further in the results section.

### LC-MS Method and data analysis

LC-MS/MS analysis was performed on a Thermo Scientific QExactive-HF (QE-HF) hybrid mass spectrometer coupled to a Evosep One LC device. The Evosep One was operated using the 200 samples per day method at a flow rate of 2 µl/min with a 7.2-minute total run time per sample [19]. The Evosep One was equipped with a 40 mm x 150 µm column from Evosep packed with Dr Maisch C18 AQ, 1.9 µm beads. The QE HF was operated in either DDA or PRM-mode using a resolution of 60,000 AGC target of 2×10^5^ for DDA and 30,000 for PRM, maximum injection time of 50 ms, and a quadrupole isolation width of 1.2 m/z. Peptides were selected for MS/MS data acquisition using an un-scheduled method and fragmented using collision energies optimised for each peptide. An electrospray voltage of 2.1 kV and capillary temperature of 300°C, with no sheath and auxiliary gas flow, was used. MaxQuant 1.6.10.43 was used for peptide and protein identification [20] using UniprotKB/Swissprot databases for Homo Sapiens containing 42437 sequences downloaded on September 2020 and a combined viral database SARS and influenza viruses including the SARS-Cov-2 virus sequences. Targeted proteomic method refinement and data analysis was performed in Skyline-daily 20.2.1.286 [21].

### Determination of LOD and LOQ

Calibration curves for LOD and LOQ determination were obtained from SARS-CoV-2 NCAP peptides in 0.1% formic acid, synthetic sputum, swab and saliva samples. To generate the calibration curves in different matrices, proteotypic peptides for NCAP were spiked into each matrix at 10 fmol/µL followed by serial dilution (2-fold) until 10 attomoles/µL was reached. For each concentration, 5 µL sample was injected on the LC-MS system in triplicates A calibration curve was created using the raw intensity values for each peptide in each matrix and a regression value (R^2^) was calculated. Limits of detection and quantitation were determined using the standard deviation of the curve and the slope values.

### Patient sample pre-processing for MS analysis

Nasopharyngeal swab samples were received in 2.5 mL of Viral Transport Medium(VTM), comprising of Anderson’s modified Hanks Balanced Salt Solution (8.0 g/L NaCl, 0.4 g/L KCl, 0.05 g/L Na_2_HPO_4_, 0.06 g/L KH_2_PO_4_, 1.0 g/L Glucose, 0.7 g/L NaHCO_3_, 0.2 g/L MgSO_4_.7H_2_O, 0.14 g/L CaCl_2_.2H_2_O) with 2% v/v heat-inactivated fetal bovine serum, 100 µg/mL gentamicin and 0.5 µg/mL amphotericin B, as recommended by the CDC [22].The virus was inactivated by heating at 80°C for 5 min and 1 mL of 20% SDS was added to the VTM containing the swab such that the total SDS concentration was ∼5 % which is compatible with the S-trap method of sample digestion. The samples were vortexed for 15 min followed by centrifugation at 1500 x g and the entire solution was collected, aliquoted and stored at -80°C for further use.

## RESULTS

A high-throughput LC-MS method should be reproducible, robust and sensitive. Towards our goal of developing a high-throughput PRM method for detection of SARS-Cov-2 peptides, we developed a fast and reproducible sample preparation method which can be used in conjunction with the PRM method. The details of the of the sample preparation and LC-MS method are listed below.

### Sample preparation for high-throughput analysis of SARS-CoV-2 samples

The sample preparation was designed to accommodate nasopharyngeal swabs which are the preferred sample collection methods in use in the clinic. In order to return clinical data quickly, we aimed to design the sample preparation method for high-throughput sample preparation, efficient virus inactivation and analysis within 2 hrs (**Figure 1**).

**Figure 1:**
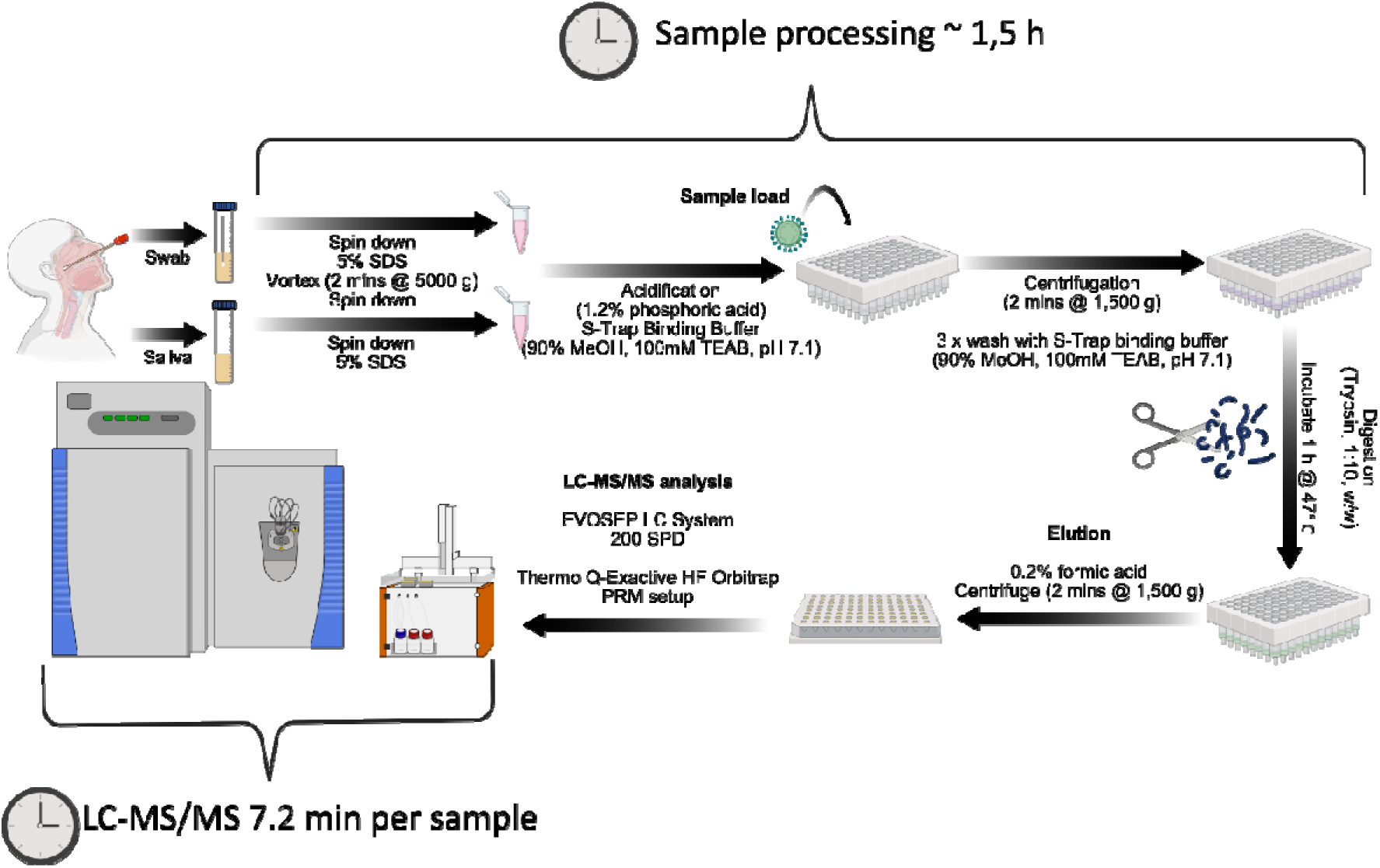
A stepwise detailed workflow for high-throughput sample processing for LC-MS based detection of SARS-CoV-2 peptides. 20% SDS was added to nasopharyngeal swab or saliva samples from patients for a final concentration of 5%. Fifty uL of the 3 mL sample were added onto 96-well S-Trap plates, where the samples were washed, digested, and eluted in a mass spectrometry compatible buffer. The eluates were directly separated on an EvoSep LC system and analysed on a QExactive HF mass spectrometer. The present method allows efficient digestion of multiple samples with various matrices under 2 hours, permitting the LC-MS and data analysis in approximately 15 mins.

Recombinant SARS-CoV-2 SPIKE and NCAP proteins were spiked at 100 pmol/mL into artificial saliva or into saliva and swab samples (stored in 3 mL viral transfer medium) taken from a healthy volunteer. As clinical samples will require virus inactivation, we tested 70 % ethanol, heat inactivation at 80°C for 5 min as well as addition of 20% SDS to a final concentration of 5% followed by heat inactivation at 80°C. After addition of ethanol or SDS, the samples were shaken for 15 mins and centrifuged for 5 mins at 1500 x g. Samples were further processed using 96-well S-trap plates as this workflow provides a fast method for digestion of proteins in most buffers without the need for precipitation, thereby avoiding sample loss and long sample preparation times. As none of the previously reported proteotypic peptides of NCAP or SPIKE contain a cysteine, we tested if reduction with TCEP and alkylation with IAA were necessary for the efficient detection of the target peptides. Digestion efficiency was assessed by LC-MS/MS analysis of tryptic peptides and was found to be unaffected even when samples were not reduced and alkylated before digestion. **(Supplementary Figure 1A)**.

In the standard S-Trap protocol peptides are eluted with two aqueous (50 mM TEAB and 0.2% formic acid) and an organic (50% MeCN, 0.2% formic acid) step. Due to the organic solvent, the samples require vacuum drying before LC-MS analysis. In order to save time, we tested if we could omit the elution in MeCN, thereby allowing straight injection of the eluted digests into the mass spectrometer. Indeed, omission of the organic elution did not affect overall intensities of SPIKE and NCAP peptides **(Supplementary Figure 1B**).

### Identification of SARS-CoV-2 proteotypic peptides for targeted analysis

Selection of appropriate, proteotypic peptides is a key step towards developing a robust and reproducible targeted LC-MS assay. In order to identify the proteotypic peptides of SARS-CoV-2 SPIKE and NCAP proteins, recombinant proteins were digested in neat (0.1% formic acid) and spiked into artificial saliva or into saliva and swab samples taken from a healthy volunteer. Digests were analysed on a Q-Exactive HF in data-dependent analysis (DDA) mode coupled to an Evosep LC -MS system, using the 60 sample per day (60 SPD) method (**Figure 2**). The DDA-based analysis of neat SPIKE and NCAP recombinant proteins revealed a list of tryptic peptides **(Supplementary Data 1)**, which provided the basis for initial screening of the target peptides. Furthermore, a QconCAT for stable isotope labelling based absolute quantification was added. This QconCAT was synthesized to include specific SPIKE and NCAP peptides based on the study by Puyvelde et al. [16]. Finally, DDA analysis of saliva samples revealed a list of consistently identified proteotypic peptides from high-abundant human proteins such as Lysozyme C (LysC) which we used as an internal control to check for sample preparation and acquisition efficiency **(Supplementary Data 2)**.

**Figure 2:**
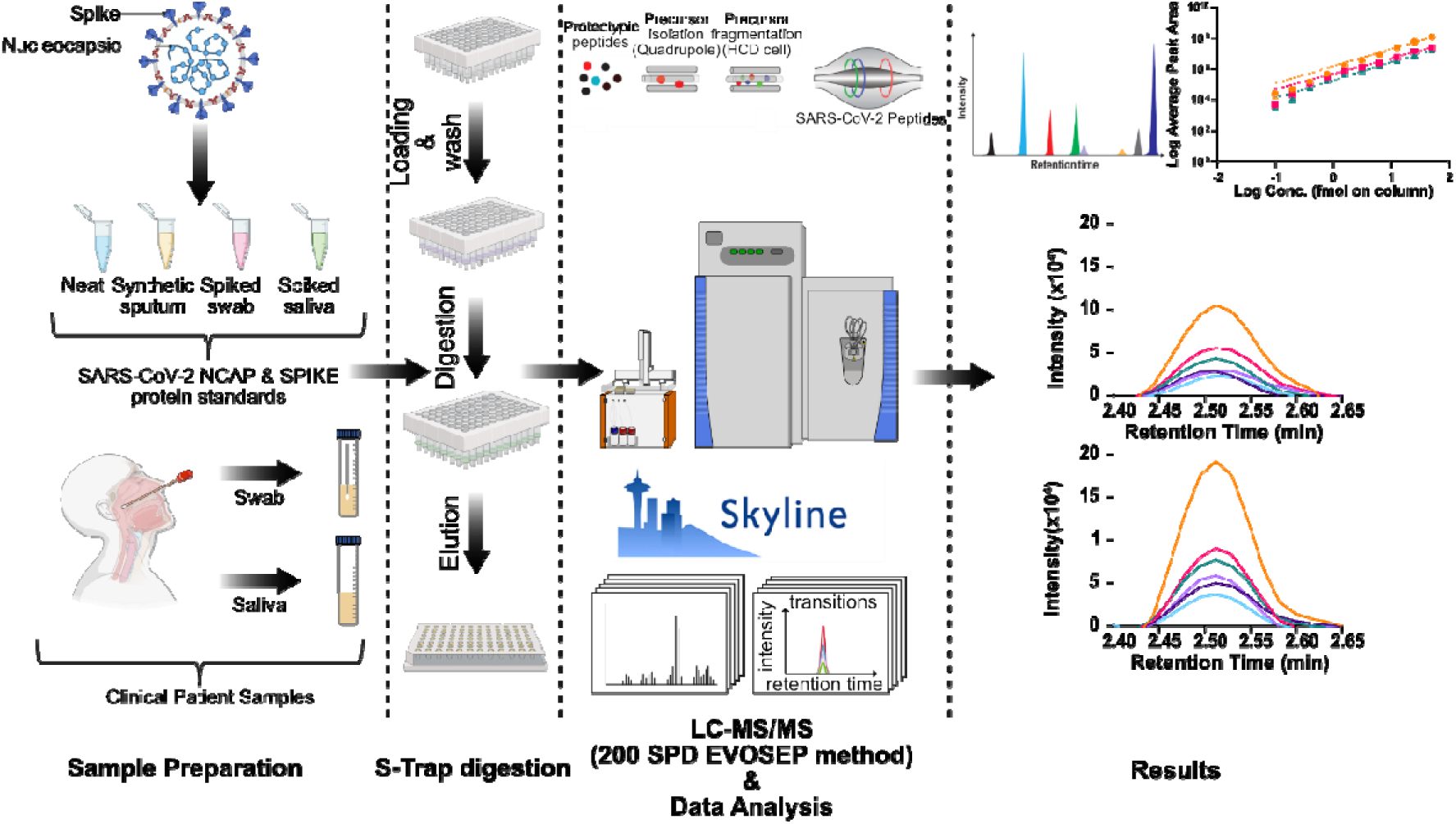
High-throughput and fast PRM-MS method for detection of SARS-CoV-2 peptides using state-of-the art LC-MS set-up. Recombinant protein standards for SPIKE and NCAP were utilized in neat, simulated sputum, saliva and nasopharyngeal swabs for developing a fast and high-throughput PRM method using 96-well S-trap sample preparation and an Evosep coupled to Thermo QE-HF mass spectrometer. Data was analysed by Skyline to assess the sensitivity of the system as well as to validate using patient swab samples.

In order to select optimal proteotypic peptides for the targeted PRM analysis, we followed published selection criteria for a targeted-based LC-MS assays such as tryptic peptide length of 8 to 25 amino acid and excluded peptides with possible modifications such as oxidation on Methionine, deamidation on Asparagine followed by Glycine, and any other possible post-translational modifications [24]. Following these criteria, an initial list of tryptic peptides was generated which were further analysed in matrices such as saliva. Of the initial 19 peptides (**Supplementary Data 3**), only five could reproducibly be detected when analysed in a background matrix of artificial and volunteer saliva. As peptides from SPIKE were inconsistently observed and their retention times varied across runs, SPIKE peptides were omitted from future analysis. SPIKE peptides are known to be heavily glycosylated, making them notoriously harder to detect [25]. Moreover, NCAP is considered to be about 3-fold more abundant in SARS-CoV-2 than SPIKE [4], increasing the sensitivity. Therefore, it was not surprising that the highest sensitivity was achieved for the NCAP peptides AYNVTQAFGR and ADETQALPQR. Additionally, as KADETQALPAR was consistently detected in all the DDA runs, it was included in the list even though it contains a missed cleavage. Human Lysozyme C, as mentioned earlier, was used as an internal control for sample preparation and to check for digestion and ionization efficiency. The final list consisted of total seven peptides including the stable isotopically labelled peptides from QconCAT which were further used for MS method development and optimization (**Table 1**).

**Table 1:** Peptides selected for the development of PRM-MS assay for detection of SARS-CoV-2 peptides.

| No | Peptide Sequence | Source | m/z, charge | Norm. Collision Energy | Type |
| --- | --- | --- | --- | --- | --- |
| 1 | AYNVTQAFGR | NCAP(SARS-CoV-2) | 563.785<sup>2+</sup> | 24 | light |
| 2 | ADETQALPQR | NCAP(SARS-CoV-2) | 564.785<sup>2+</sup> | 23 | light |
| 3 | KADETQALPQR | NCAP(SARS-CoV-2) | 419.557<sup>3+</sup> | 20 | light |
| 4 | AYNVTQAFGR | QconCAT | 571.263<sup>2+</sup> | 24 | heavy |
| 5 | ADETQALPQR | QconCAT | 572.263<sup>2+</sup> | 23 | heavy |
| 6 | KADETQALPQR | QconCAT | 425.207<sup>2+</sup> | 20 | heavy |
| 7 | STDYGIFQINS | LYS-C (*Homo sapiens*) | 700.843<sup>2+</sup> | 25 | light |

### PRM-MS method development and optimization

A PRM method was created on the QE-HF using the 200 samples per day (200 SPD) short gradient LC method integrated on the Evosep One. The Evosep One is specifically designed to run samples back-to-back without carryover due to the tip-based sample injection system, which serves as a single-use trap column thereby increasing the overall life of the column, making the system well-suited for a high-throughput method [26]. The PRM method was further optimized for collision energies (**Supplementary Figure 2)** for the individually selected peptides along with their corresponding SIL peptides emerging from the QconCAT. The QconCAT sequence information can be found in (**Supplementary Data 4**). The normalized collision energies (NCE) for the targeted peptides were optimized within the range of 20-29 for all the peptides. The optimal collision energies for individual peptides were selected based on the intensities of their product ions using Skyline. Details of the seven peptides and collision energies used for the final PRM assay are listed in **Table 1**.

### Assessing the sensitivity and robustness of the PRM method

The next step in establishing the high throughput PRM method involved its assessment for sensitivity and specificity. Calibration curves of the target peptides in neat, artificial saliva as well as real saliva and swab samples were generated using the tryptic digests for recombinant protein standards for NCAP. Recombinant tryptic peptides in 0.1% formic acid (“neat”) **(Figure 3A)**, spiked swab **(Figure 3B)**, spiked oral fluid **(Figure 3C)** and spiked saliva **(Figure 3D)** were injected in a dilution series from 50 femtomoles to 50 attomoles was on column. The limit of detection (LOD) for NCAP peptides was found to be as low as 170 amol on column, while the limit of quantitation (LOQ) was found to be 850 amol when injected neat (**Table 2**). The calibration curves for the logged intensities also depict the linearity of the curves as seen in **(Supplementary Figure 3)**. The LOD and LOQ were calculated using the formulae 3*Sa/b and 10*Sa/b respectively, where Sa is standard deviation of the calibration curve response and b is the slope of the curve. The LOD and LOQ decreased to 0.63 fmol and 3.24 fmol on column, respectively, for the same peptide when injected as a spike-in in the real saliva and swab samples. The drop in sensitivity was expected when peptide standards were injected with saliva sample due to the matrix effect contributed by other endogenous high abundant salivary proteins such as Lysozyme C and Albumin. The high abundance of these proteins contributes towards signal suppression of the target peptides resulting in the overall drop in sensitivity in the saliva and swab samples.

**Figure 3:**
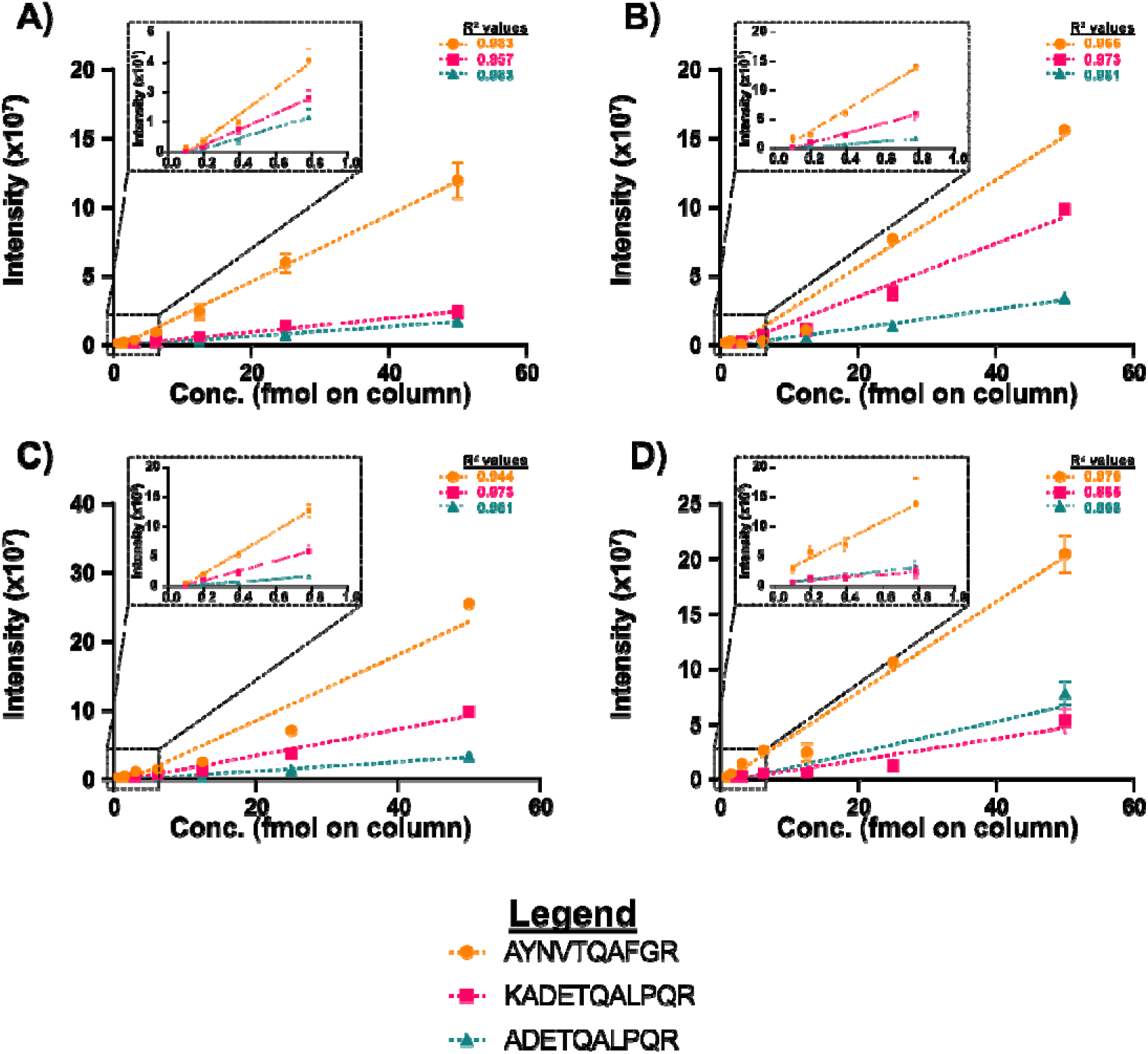
Calibration curves for estimating the limits of detection (LOD) and quantitation (LOQ). Tryptic peptides generated from recombinant SARS-CoV-2 NCAP standard protein were injected on the LC-MS system to generate calibration curves starting from 50 fmol on column and diluted 2-fold until 48 amol was achieved either in 0.1% formic acid as depicted in figure A. Figures B, C and D represent the drop in sensitivity of the LOD and LOQ when injected as a spike-in in a swab, oral fluid or saliva sample respectively. The sensitivity of the system was assessed based on the three peptides AYNVTQAFGR, ADETQALPQR and KADETQALPQR selected for final PRM method. The figures in the inset are the zoomed areas of the lower concentration data points.

**Table 2:** Limits of detection and quantitation calculated for NCAP peptides on the existing LC-MS system in various matrices are represented below.

| Protein | Matrix | Peptide | LOD (fmol on column) | LOQ (fmol on column) |
| --- | --- | --- | --- | --- |
| NCAP | 0.1% Formic acid | AYNVTQAFGR | 0.17 | 0.85 |
|  |  | ADETQALPQR | 0.37 | 1.1 |
|  |  | KADETQALPQR | 0.26 | 1.3 |
|  | Artificial sputum | AYNVTQAFGR | 1.6 | 8.0 |
|  |  | ADETQALPQR | 0.66 | 3.3 |
|  |  | KADETQALPQR | 6.3 | 32.1 |
|  | Spiked Saliva | AYNVTQAFGR | 2.6 | 12.8 |
|  |  | ADETQALPQR | 4.7 | 23.6 |
|  |  | KADETQALPQR | 7.3 | 36.4 |
|  | Spiked Swab | AYNVTQAFGR | 1.5 | 7.2 |
|  |  | ADETQALPQR | 3.0 | 14.8 |
|  |  | KADETQALPQR | 4.4 | 21.9 |

The existing PRM assay system needed to not only be sensitive but also robust to carryout high-throughput analysis of many samples. To test the robustness of the LC-MS system a continuous 200 sample test was performed with a saliva sample spiked-in with NCAP peptides. The peptide AYNVTQAFGR was monitored for its product ion areas and retention time. The test revealed that the system performed consistently without a notable drop in sensitivity across all 200 samples, solidifying the stability of the system to handle such a load (**Figure 4A and 4B)**.

**Figure 4:**
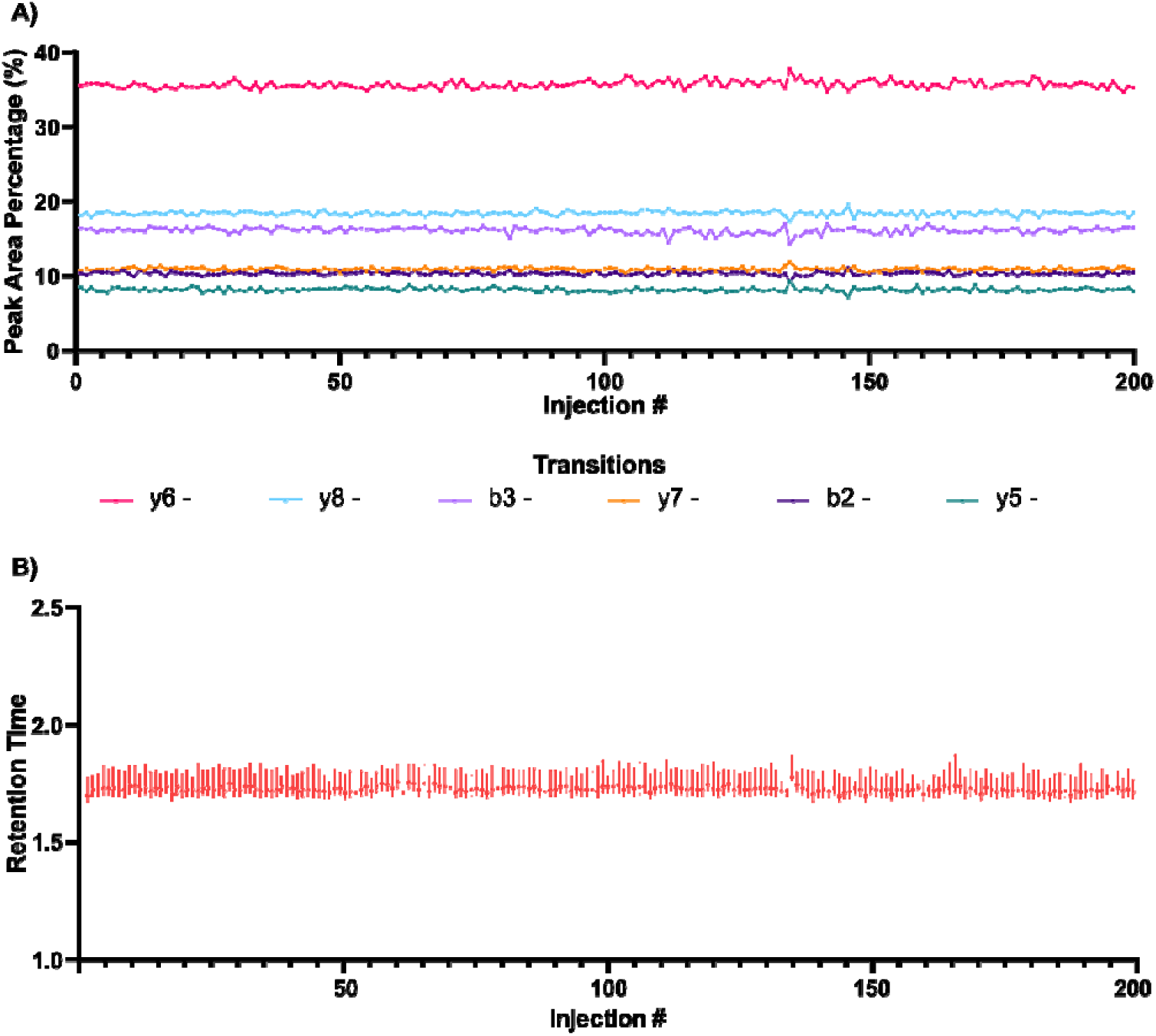
Testing the robustness of the LC-MS assay. To test the stability and robustness of the existing PRM method, a stress test with 200 consecutive injections of saliva spiked with neat NCAP peptides was conducted where figure A depicts overall consistent peak area percentage for all the 6 transitions for the NCAP peptide AYNTQAFGR. Figure B depicts the intensity and retention time variation of the same peptide over 200 injections.

### Validation of PRM Assay using 50 patient samples

The final PRM method was validated using 25 positive and 25 negative patient samples. The samples had been analysed by either RT-PCR, SAMBA II or LumiraDx. RT-PCR provided a Cycle Time (Ct) value and any Ct <40 was considered as positive. The Simple AMplification-Based Assay (SAMBA) II nucleic acid testing system (SAMBA II) finds traces of viral genetic material and amplifies it a billion times [28] and LumiraDx is a microfluidic immunofluorescence assay for qualitative detection of nucleocapsid protein antigen [29]. The latter two methods do not provide a quantitation of viral load.

The samples were processed according to the optimized sample preparation protocol. BCA estimation of the samples revealed that the total concentration of the protein in the extraction buffer ranged between 3-5 mg/mL. Out of the total sample extracted from swab in 3 mL of VTM with 5% SDS buffer, only 50 μL sample (∼200 μg) was taken for processing. ∼10 μg sample was injected on column, where concentration of QconCAT in the sample was 10 fmol on column. Thus, ∼1/1200^th^ of the total sample was used for detection of the SARS-CoV-2 peptides using the present method.

The samples were attributed as positive only if they passed the following criteria: (i) At least two out of maximum six fragment ion transitions were detectable and (ii) The retention times matched that of the heavy QconCAT counterpart of the same peptide. The peptides that were assigned as positive were then used for calculation of the probable absolute concentration from the total H/L ratios calculated by Skyline software.

Our PRM method was able to detect nine positive cases, three shown here and the other six in (**Supplementary Figure 4**), out of the 25 positive samples correctly (**Figure 5)**, suggesting that patients with high viral load can be detected with this method. LUMIRA, qPCR and SAMBA II results for all 50 patient samples are given in (**Supplementary Data 5**).

**Figure 5:**
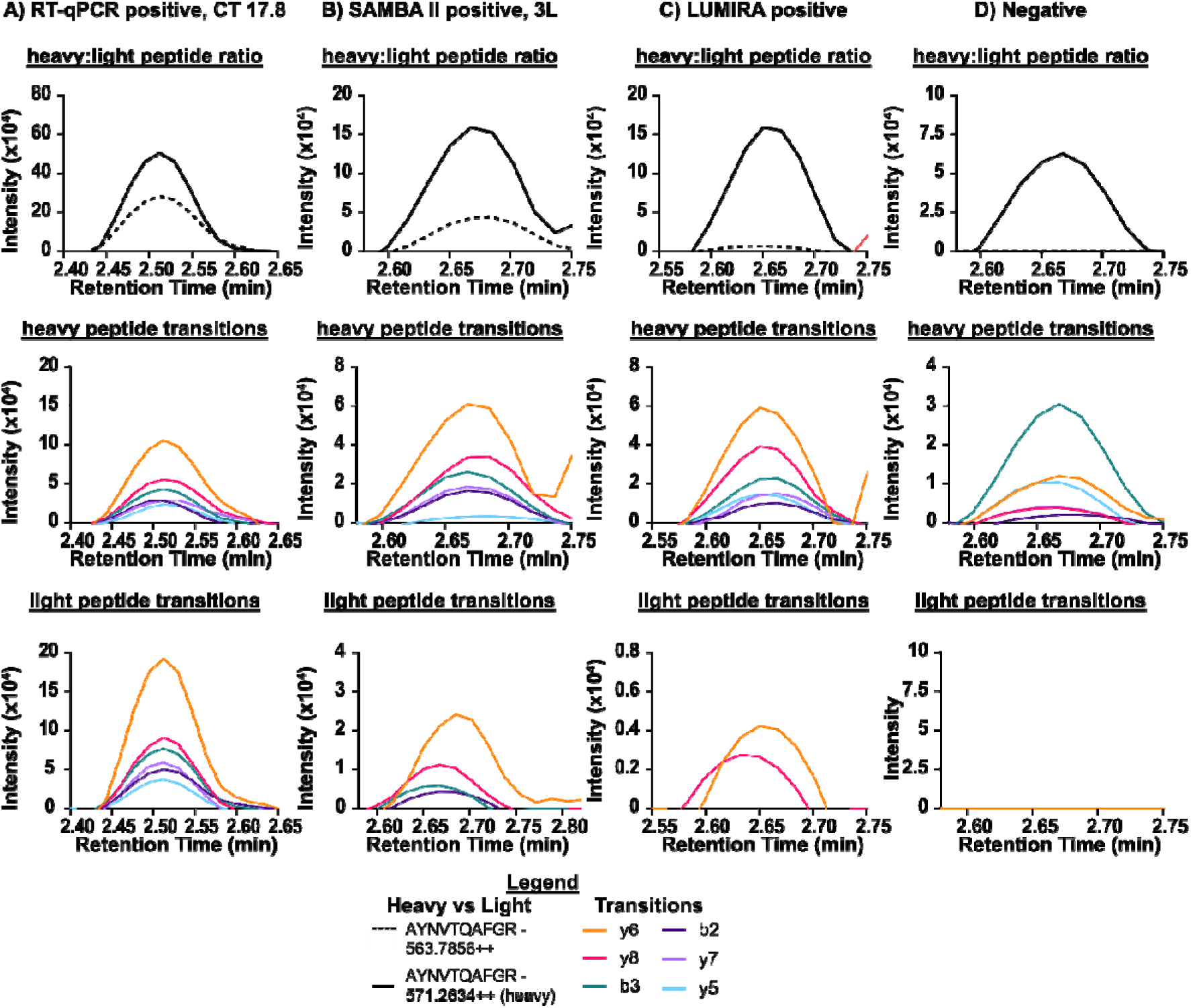
PRM transitions detected in patient samples. **A**) Sample of a positive patient where the viral particles were detected by RT-qPCR with a CT value of 17.8. The concentration of the viral peptide AYNTQAFGR was estimated to be ∼25 fmol (corresponds to 7.5 pmol in the whole swab sample) by calculating the heavy to light ratio (H/L). **B)** and **C)** are patient samples deemed positive with SAMBA II and Lumira, respectively, which were also confirmed with our method as positives. **D)** is a negative patient sample.

## Discussion

The presented MS-based assay aims to directly detect viral peptides in nasopharyngeal swab samples with faster processing times in a high-throughput manner. The detection is pursued from miniscule amounts of total samples unlike the nucleic acid-based strategies which utilise amplification. This is one of the major reasons attributing to the lower sensitivity of the present MS-based targeted assay compared to nucleic acid amplification-based methods such as PCR and SAMBA. While Lumira does not require amplification, it requires antibodies which can be non-specific and can lead to false positives. In order to achieve more sensitivity higher sample volume needs to be injected onto the system, which poses a challenge when using a nano-flow LC system.

Additionally, to provide a boost in sensitivity pre-enrichment methods such as SISCAPA [30], aptamers [31] or bead-based enrichments as utilized by Renuse et.al [15] could be incorporated in the sample preparation workflow, as the viral peptides have very low abundance when measured against a background of human saliva samples rich with high-abundant proteins like Lys C and Amylase. The other reason that contributes towards lowered detection sensitivity of the PRM assay could be assigned to the way the samples were collected and stored. In many of the samples, the traces of food particles and phlegm from individual patients were found, which made sample processing problematic. This highlights the fact that sample collection and storage for MS-based assays should be standardized, such as making sure the nasopharyngeal swab samples are taken by qualified staff, making sure the swabs touch only the back of the throat and the base of the tonsils avoiding any contamination. Also, efforts should be taken towards immediate processing of the samples after collection or stored at -20°C to reduce any protein degradation.

Stable isotope standards help in achieving absolute quantification [32]. One such standard is QconCAT (Quantification conCATemer), which is an artificially synthesized protein, generated by concatenation of proteotypic peptides. A heavy labelled QConCAT internal standard enables assessing sampling quality, sample preparation efficacy, instrument robustness, and absolute quantification [16]. Heavy labelled QconCAT synthesized specifically to contain NCAP and SPIKE proteins for SARS-CoV-2 along with other host proteins enabled the absolute quantification of SARS-CoV-2 peptides. Among the patient samples, one sample had been analysed by RT-PCR with a Ct value of 17.8, which indicates a severe viral load. We calculated the amount for the viral NCAP peptide (AYNTQAFGR) to be 25 fmol on column. Assuming that each viral particle contains 300-350 NCAP molecules [33], this indicates that the swab contained approximately 5.25 × 10^8^ viruses. This is in the right order of magnitude with the theoretical calculations conducted by Puyvelde *et al* [16], where a correlation of number of viral particles from measured Ct values of plasmids was attempted. They estimated that a Ct value of 16 would amount to ∼20 fmol NCAP/10 μL sample corresponding to 1.26 × 10^10^ NCAP copies or 4.2 × 10^8^ viral particles (one RNA per virus) per 10 μL sample. This shows good correlation of our PRM method with existing methods, as well as enables calculation of viral particles from absolute concentrations of viral proteins. Indeed, more rigorous studies with a greater number of patient samples with standardized sampling protocols are needed to establish a strong peptide concentration-viral particles correlation baseline. Nonetheless, the quantification of these peptides in the nasopharyngeal samples is a step in the right direction to assess not only the prognostic potential of the method but also determining the concentration range of the protein in correlation with the severity of the SARS-CoV-2 disease, which may prove helpful in designing treatments for disease management and thereby reducing complications and hopefully fatalities.

### Future Perspectives

The global response to the Covid-19 pandemic has come a long way since its inception in late 2019. Lately, the death toll and the number of SARS-CoV-2 variants arising with the on-going pandemic seem to be under control with the introduction of several vaccines, yet complete global immunization against Covid-19 is still far away. With the world starting to turn slowly towards normality, rigorous testing remains of prime importance until Covid-19 is completely eradicated which puts a burden on existing molecular testing methods. The present method could serve as an alternative for fast detection, where such mass spectrometric and chromatographic capabilities are available. With the little requirements for development of an MS-based testing method, it presents with a relative possibility of adapting the existing LC-MS method for detection of the existing and newly emerging SARS-CoV-2 variants as well as multiplexing with other respiratory viruses such as influenza.

The sensitivity of the present method has room for improvement by incorporating enrichment methods such as SISCAPA and immunoaffinity in the sample preparation. Additionally, applying automated sample handling to the sample preparation workflow may further reduce the variability and improve the turnover times.

## Data Availability

All data produced in the present study are available upon reasonable request to the authors

## Acknowledgements

We would like to thank Sam Dhesi and Luke Wainwright from the Department of Health and Social Care as well as Perdita Barran for their support. We would like to thank all members of the Laboratory for Biomedical Mass Spectrometry for their support during this project. This work was funded by the UK Department of Health and Social Care. AN is funded by a Marie Sklodowska Curie ITN studentship within the European Union’s Horizon 2020 research and innovation programme under the Marie Skłodowska-Curie grant agreement No. 861316. FS is funded by a BBSRC iCASE studentship (2306774). AG, TH and MT are funded through a Wellcome Investigator Award (215542/Z/19/Z)

## Author contributions

AG, AN, TH, FS, AP performed experiments and data analysis. YT provided access to patient samples. AG, AN and MT wrote the manuscript with input of all authors.

