## Supplementary Figures for "A fast and sensitive absolute quantification assay for the detection of SARS-CoV□2 peptides using Parallel Reaction Monitoring Mass Spectrometry"

**1A)**

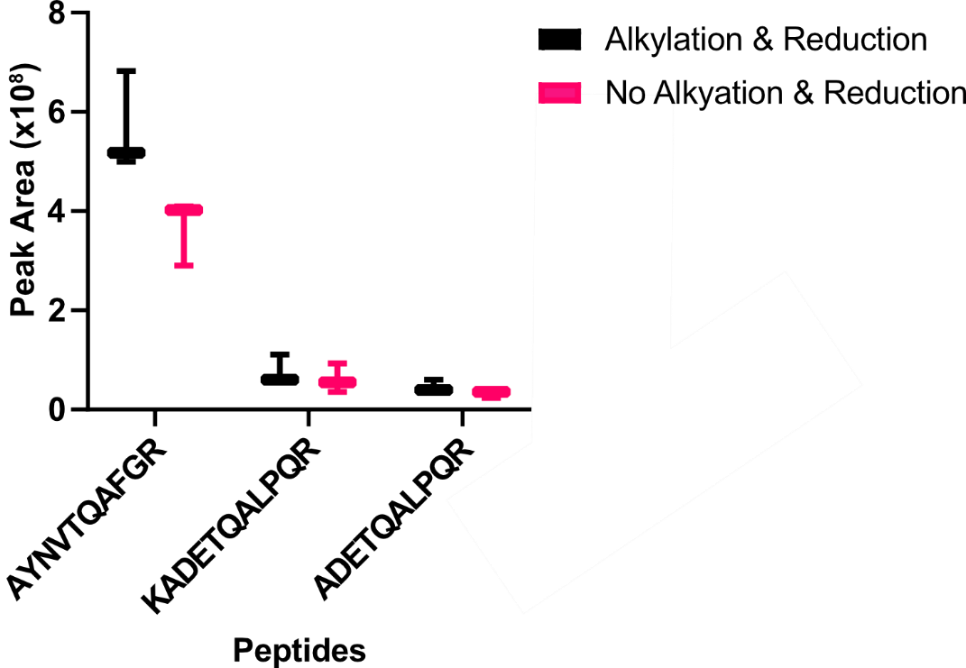

**1B)**

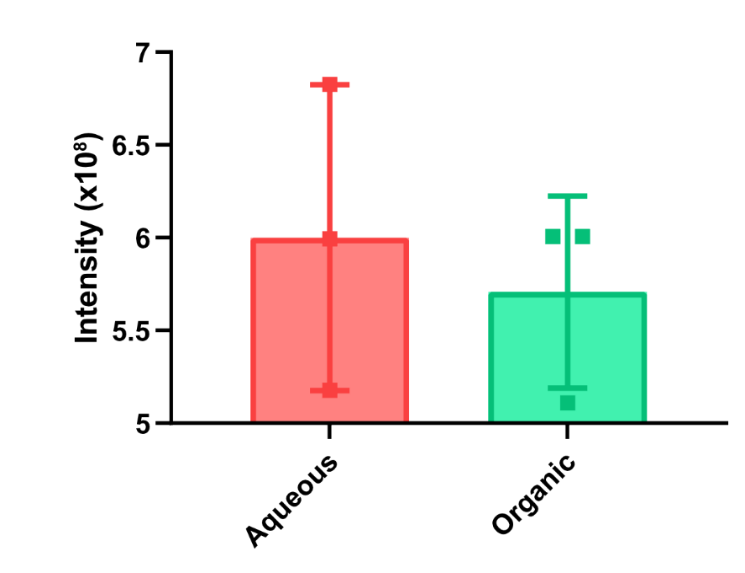

**Supplementary Figure 1: Optimising digestion of NCAP protein. (A)** Digestion efficiency of the NCAP peptides was checked with and without alkylation and reduction showing that reduction and alkylation was not necessary. (B) Elution from the S-trap with or without organic solvent shows the slight improvement of the peptide recovery when only aqueous elution (50mM TEAB and 0.1% Formic acid) was used compared to organic elution (50mM TEAB, 0.1% Formic acid and 50% acetonitrile; 0.1% Formic acid).

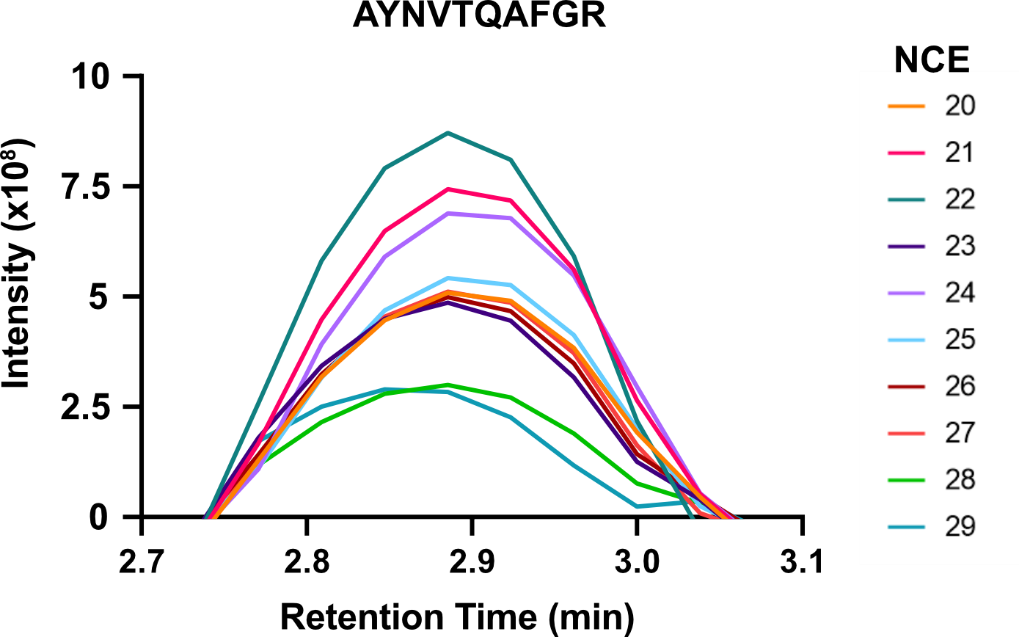

**Supplementary Figure 2: Optimisation of collision energies for NCAP peptide transitions.** Collision energies were optimized for all targeted peptides and are included in the final PRM method. The figure above shows all the collision energies used for optimization for the peptide AYNVTQAFGR, where NCE 22 was selected in the final method.

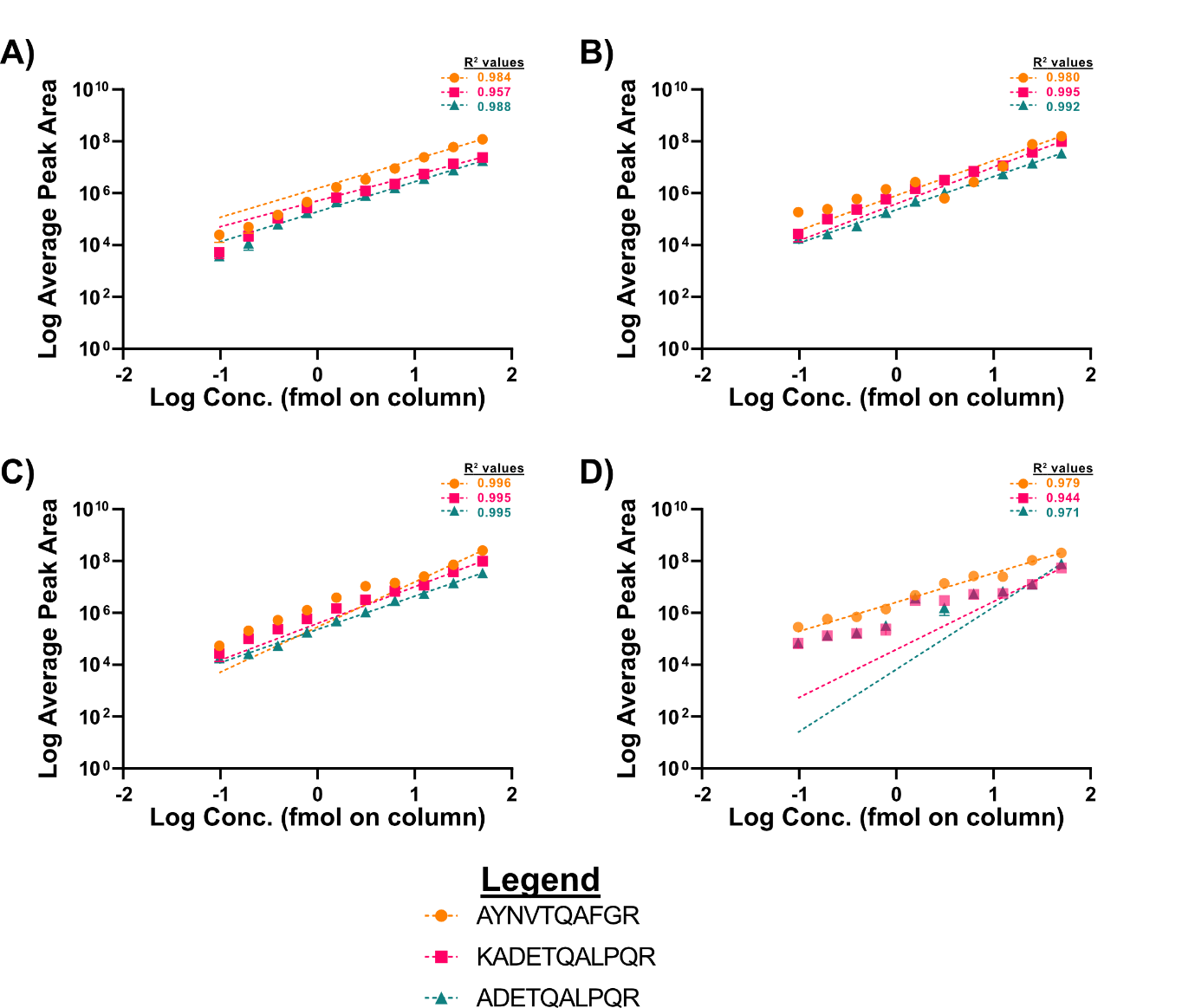

**Supplementary Figure 3:** Calibration curves with logged intensity and concentration of the 2-fold dilution of NCAP peptides in A) 0.1% Formic acid, B) Spiked swab, C) Spiked oral fluid and D) spiked saliva. The curves show good linearity.

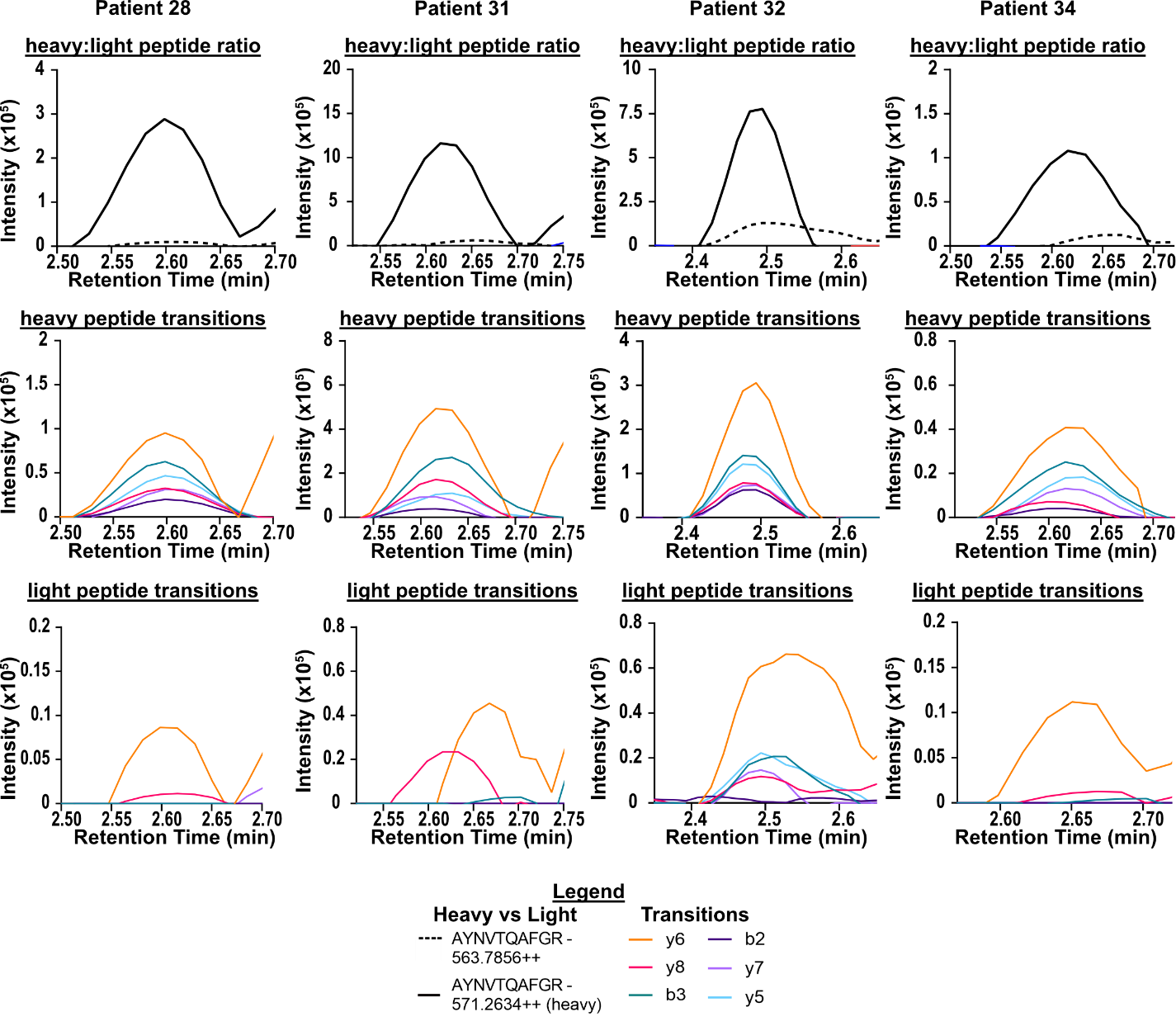

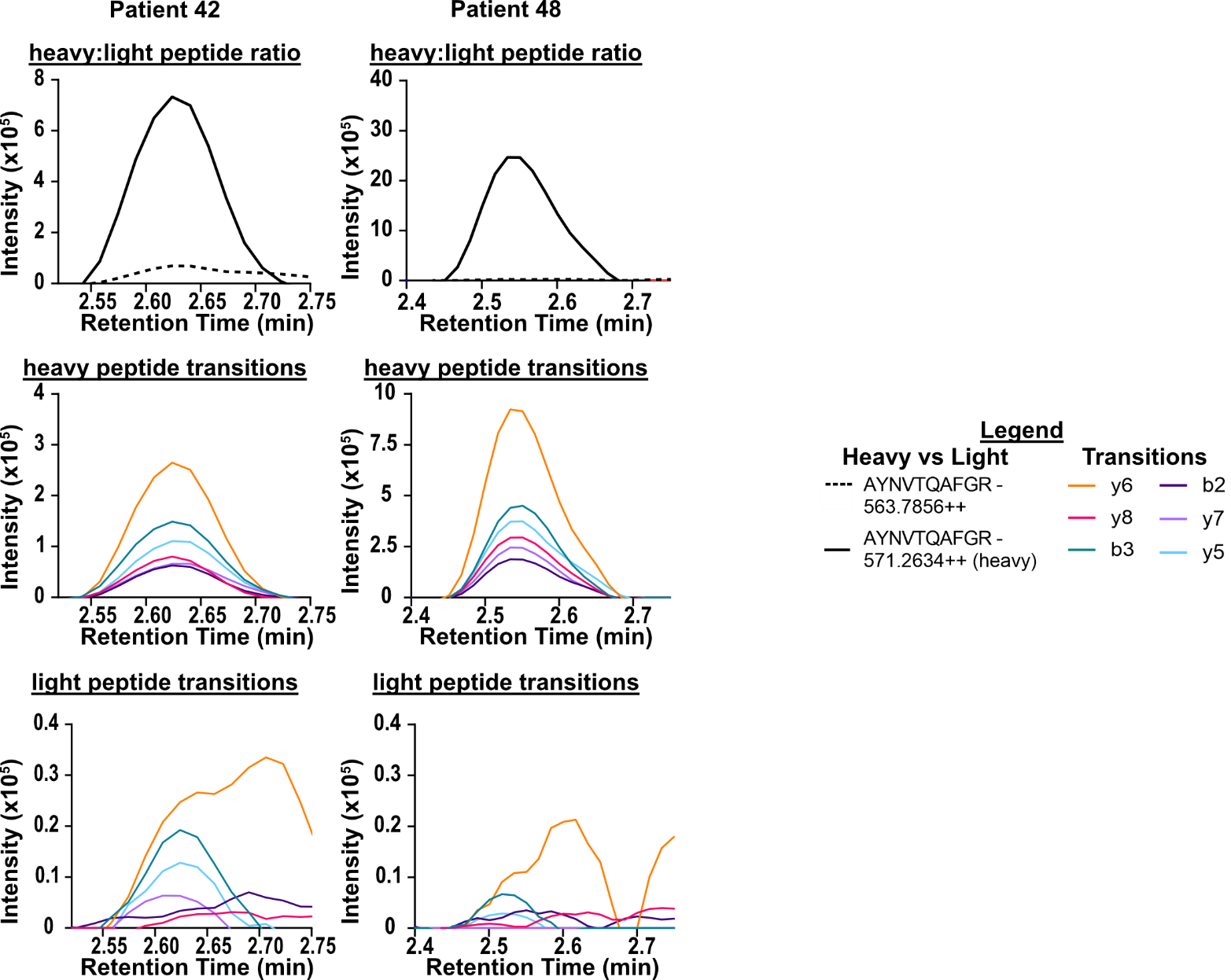

**Supplementary Figure 4: PRM transitions detected in patient samples.** Remaining six out of total nine samples detected as positive with the existing PRM method.
